# Disparities in Excess Deaths from the COVID-19 Pandemic Among Migrant Workers in Kuwait

**DOI:** 10.1101/2021.03.25.21254360

**Authors:** Barrak Alahmad, Dawoud AlMekhled, Ayah Odeh, Janvier Gasana

## Abstract

**Background:** The actual human cost of the pandemic cannot be viewed through the COVID-19 mortality rates alone. Especially when the pandemic is widening the existing health disparities among different subpopulations within the same society. In Kuwait, migrant workers were already disproportionately impacted by COVID-19 and its unintended consequences.

**Objective:** To estimate the excess deaths in the pandemic year of 2020 among the Kuwaitis and non-Kuwaiti migrants.

**Methods:** We analyzed publicly available retrospective data on total annual mortality historically (2005 to 2019) and in 2020. We fitted a quasi-poisson generalized linear model adjusted for yearly trend and nationality to estimate the expected deaths in 2020 in the absence of the pandemic. We calculated excess deaths as the difference between observed and expected mortality for the year of the pandemic in both Kuwaitis and non-Kuwaitis.

**Results:** In the absence of the pandemic, we expect the total mortality in Kuwait to be 6629 (95% CI: 6472 to 6789) deaths. However, the observed total mortality in 2020 was 9975 deaths; about 3346 (3186 to 3503) more deaths above the historical trend. Deaths among migrant workers would have been approximately 71.9% (67.8 to 76.0) lower in the absence of the pandemic. On the other hand, deaths among Kuwaitis would have been 32.4% (29.3 to 35.6) lower if the country had not had the pandemic.

**Conclusion:** The mortality burden of the COVID-19 pandemic is substantially higher than what the official tally might suggest. Systematically disadvantaged migrant workers shouldered a larger burden of deaths in the pandemic year. Public health interventions must consider structural and societal determinants that give rise to the health disparities seen among migrant workers.

## Introduction

The coronavirus disease 2019 (COVID-19) pandemic has triggered a mass disruption to social, economic, and environmental modes of operation. Of utmost concern to population health and wellbeing is the disruption to the health system as caused by the pandemic (1). Governments worldwide have put efforts to mitigate the burden of the pandemic on the healthcare system by reducing access to it; through cutting down routine testing, cancelling elective surgeries and the follow-up of chronic conditions (1,2). Henceforth, the human cost of the pandemic cannot be viewed through the COVID-19 mortality rates alone as disruptions to healthcare access caused by the pandemic are likely to result in greater mortality. Furthermore, because of the differences in death reporting protocols, using COVID-19 reported deaths makes the comparison between countries difficult(3).

In order to assess the overall impact of a pandemic, excess death must be carefully examined (3,4). Excess death is the difference between the number of deaths from all causes observed during a public health crisis and the expected number of deaths in its absence or in otherwise ‘normal’ conditions. This provides an efficient mortality surveillance strategy accounting for the indirect burden of disease caused by disruptions to access and use of healthcare (4). Such estimate overcomes the variability between different countries in how they report COVID-19 deaths as well as allowing for a clearer indication of the impact of the pandemic. Excess deaths have been previously used to assess the human costs of prior pandemics and to estimate the impact of climate change on mortality (5–9).

Ethnic and racial disparities have existed in healthcare access and utilization prior to the COVID-19 pandemic (1,10). In the US alone, the Black community has experienced lower standards of health, lower life expectancies, and higher mortalities compared to their white counterparts (11,12). Attributable to the social determinants of health, marginalized communities’ differential access to education, housing, healthcare and employment reduces their quality of overall health leading to a lower quality of life.

Kuwait has been severely affected by COVID-19. Due to lockdowns and curfews the country has witnessed a disruption to healthcare access, delivery, and provision. Kuwait has a population of 4.7 million, with a demographic profile consisting of approximately two-thirds non-Kuwaitis (13). The majority of non-Kuwaitis (and similarly non-citizens in other Gulf countries) are migrant workers employed in low-skilled sectors and domestic work with precarious working conditions and irregular healthcare access. Non-Kuwaitis are more likely to work in occupations that increase the risk of transmission, live in poor or cramped housing, and face language and cultural barriers (14). Previous studies in Kuwait have highlighted the disparities in COVID-19 exposure risk and adverse health outcomes (14–16). However, to date there has been no study assessing the excess deaths in Kuwait as well as between Kuwaitis and non-Kuwaitis attributable to the COVID-19 pandemic. This study aims to estimate the excess deaths in the pandemic year of 2020 among the Kuwaiti and non-Kuwaiti subpopulations.

## Methods

### Data Sources

Data from Kuwait was collated from official sources that are made publicly available by the Kuwaiti Government. Annual population estimates by nationality from 2005 to 2020 was obtained from the Public Authority for Civil Information (accessible: https://www.paci.gov.kw/stat/TimeSeries.aspx). Similarly, deaths by nationality in 2005 and 2020 were obtained from the same source. In an additional analysis, we used historical mortality data between 2001 and 2018 by nationality from annual mortality reports prepared by the government’s Central Statistical Bureau (accessible: https://csb.gov.kw/Pages/Statistics?ID=10&ParentCatID=1). The compiled data that supports this analysis is available in the supplemental material (**Supplemental Raw Data**).

For a global comparison dataset of excess deaths, we used the COVID-19 excess deaths data from ‘Our World In Data’ – a project of the Global Change Data Lab, a registered charity in England and Wales that is run by Oxford Marin School, University of Oxford (accessible: https://ourworldindata.org/excess-mortality-covid)(17). We summed weekly or monthly observed and expected deaths from countries that provided complete datasets.

### Statistical Analysis

We calculated an unadjusted death rate as number of deaths divided by the population estimate per 1,000 individuals per year. We used two methods to calculate expected deaths in 2020. First, we fitted a generalized linear regression for rates of annual deaths from 2001 to 2019 with a quasi-poisson distribution to account for overdispersion. We included two independent variables: linear trend for year, and a dummy variable for nationality (Kuwaiti vs. Non-Kuwaiti). Additionally, we included an offset term log(population) to model the rate rather than counts and account for changes in population dynamics over the years. We then used the model to predict the expected 2020 deaths with 95% confidence intervals (CI) for Kuwaitis and non-Kuwaitis. In a sensitivity analysis, to check for non-linearity of yearly trend of mortality rates, we fitted a generalized additive model with penalized splines for year.

Secondly, we also used simple rolling average method to calculate the expected 2020 deaths. We averaged deaths in the last five years (2015 to 2019) for Kuwaitis and non-Kuwaitis. We calculated the uncertainty around the averaging estimate (95% CI) by adding and subtracting the estimate from 1.96 times the square root of the estimate(18).

Excess deaths were calculated as observed deaths minus expected deaths in 2020 for each subpopulation. Percent increase in excess deaths was calculated by subtracting expected from observed deaths and then dividing by expected deaths. Similar approach was carried out for upper and lower bounds of expected deaths from the two methods.

## Results

The number of deaths and death rates per 1,000 from 2015 to 2020 are presented in **Table 1**. Overall, 9975 deaths were reported in 2020, with an overall rate of 2.1 deaths per 1,000 population; a marked increase compared to the previous five years. Similarly, when examined by nationality, the total number of deaths among Kuwaitis in 2020 was 4756 with a rate of 3.3 deaths per 1,000 population, while non-Kuwait deaths totaled to 5219 deaths with a rate of 1.6 per 1,000 population.

**Table 1:** Total mortality counts and death rates in Kuwait from 2015 to 2020.

| Year | Total |  | Kuwaitis |  | Non-Kuwaitis |  |
| --- | --- | --- | --- | --- | --- | --- |
|  | Deaths | Rate per 1,000 | Deaths | Rate per 1,000 | Deaths | Rate per 1,000 |
| 2015 | 6099 | 1.4 | 3261 | 2.5 | 2838 | 1.0 |
| 2016 | 6248 | 1.4 | 3439 | 2.6 | 2809 | 0.9 |
| 2017 | 6356 | 1.4 | 3432 | 2.5 | 2924 | 0.9 |
| 2018 | 6374 | 1.4 | 3433 | 2.4 | 2941 | 0.9 |
| 2019 | 6896 | 1.4 | 3687 | 2.6 | 3209 | 1.0 |
| 2020 | 9975 | 2.1 | 4756 | 3.3 | 5219 | 1.6 |

Source: Public Authority for Civil Information, Government of Kuwait.

Over the entire historical period of annual mortality, there was a general downward trend. The pandemic year of 2020 clearly exhibits a large increase in mortality rate compared to the previous years. This was seen among non-Kuwaitis, increasing from 1.0 to 1.6, and among Kuwaitis, which increased from 2.6 to 3.3 deaths per 1,000 population (**Figure 1**).

**Figure 1.**
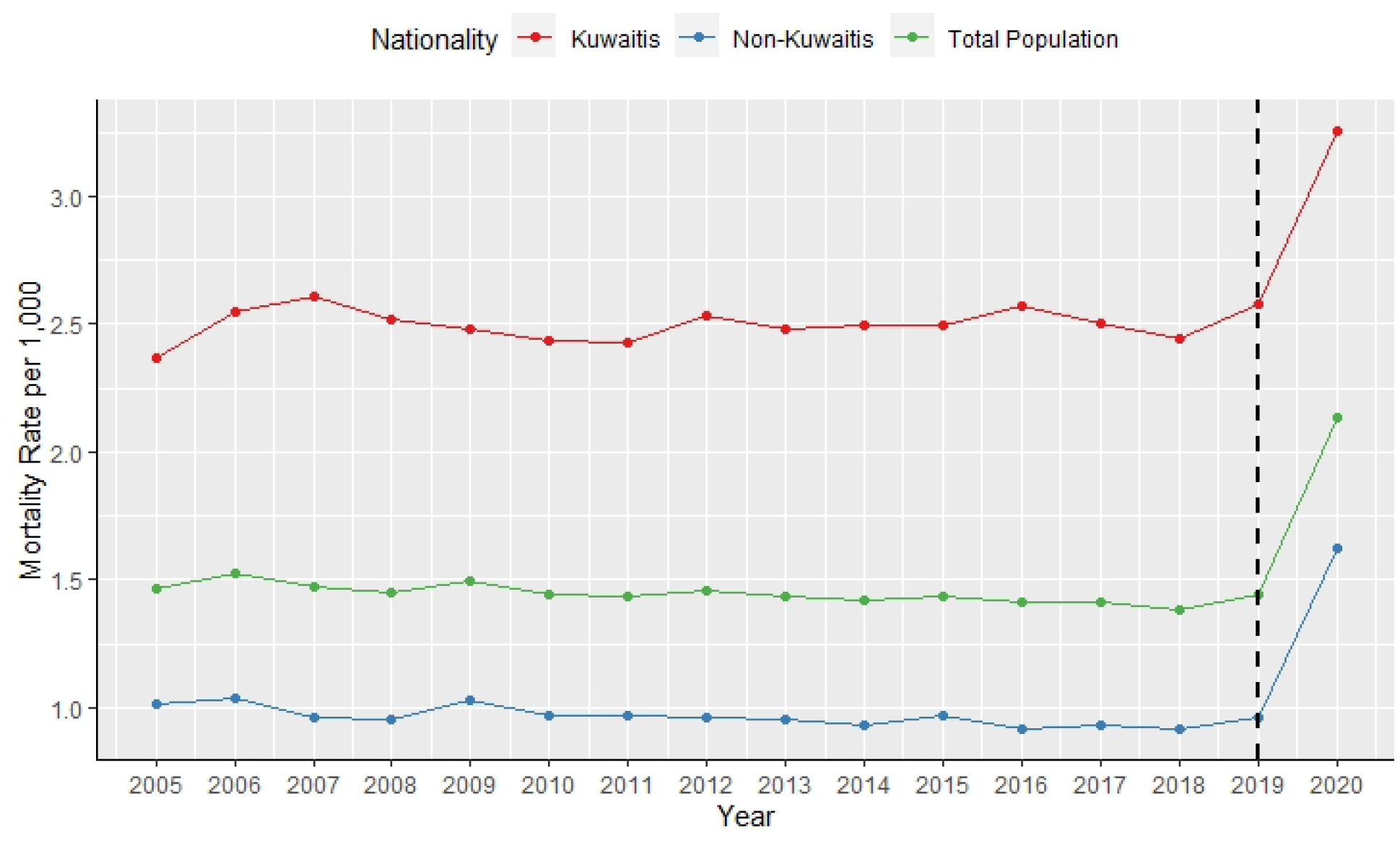
Historical mortality rates in Kuwait per 1,000 population from 2005 to 2020.

Regression analysis showed that expected deaths were significantly lower than observed deaths in 2020, with a total of 3346 (95% CI: 3186 to 3503) excess deaths occurring, a 50.5% (95% CI: 46.9 to 54.1) increase. An estimated 32.4% (95% CI: 29.3 to 35.6) increase in excess deaths was seen among Kuwaitis while 71.9% (95%CI: 67.8 to 76.0) were seen among non-Kuwaitis. Penalized spline on yearly trend did not suggest evidence of non-linearity (degrees of freedom = 1). Simple average estimates showed similar results but slightly higher than the regression analysis, with 3580 (95% CI: 3424 to 3737) more individuals than expected dying, accounting for 56.0% increase (95%CI: 52.3 to 59.9) as shown in **Table 2**. Using the Central Statistical Bureau historical mortality, the results provided similar pattern, yet the estimates were slightly lower (**Supplemental Table A**).

**Table 2:** Estimates of expected and excess deaths in 2020 stratified by nationality.

|  | **Total** | **Kuwaitis** | **Non-Kuwaitis** |
| --- | --- | --- | --- |
| **Observed Deaths in 2020<sup>a</sup>** | 9975 | 4756 | 5219 |
| **Regression Estimate:** |  |  |  |
| *Expected Deaths (95% CI)* | 6629 (6472 to 6789) | 3592 (3508 to 3678) | 3036 (2964 to 3111) |
| *Excess Deaths (95% CI)* | 3346 (3186 to 3503) | 1164 (1078 to 1247) | 2183 (2108 to 2255) |
| *Excess Deaths % (95% CI)* | 50.5 (46.9 to 54.1) | 32.4 (29.3 to 35.6) | 71.9 (67.8 to 76.0) |
| **Simple Averaging Estimate:** |  |  |  |
| *Expected Deaths (95% CI)* | 6395 (6238 to 6551) | 3450 (3335 to 3566) | 2944 (2838 to 3051) |
| *Excess Deaths (95% CI)* | 3580 (3424 to 3737) | 1306 (1190 to 1420) | 2275 (2168 to 2381) |
| *Excess Deaths % (95% CI)* | 56.0 (52.3 to 59.9) | 37.8 (33.9 to 42.6) | 77.2 (71.1 to 83.9) |

<sup>a</sup> Source: Public Authority for Civil Information, Government of Kuwait.

## Discussion

With the pandemic widening the existing health disparities among different subpopulations within the same society, we saw migrant workers shouldering a larger burden of mortality. There were more than two thousand overall excess deaths among non-Kuwaitis translating into more than 71% increase in expected mortality. In contrast, the Kuwaitis also experienced a deadly year, yet the percentage increase was nearly 32%, not as dramatic as non-Kuwaitis. That is, nearly 40 percentage points difference between the two subpopulations in an absolute scale.

The non-Kuwaiti population is comprised vastly by migrant workers who are in working age and hence relatively younger than Kuwaiti adults (13). The worker population is generally considered healthier than the comparison or standard population(19). This can be clearly seen as the historical death rates among the Kuwaitis were nearly 2.5-fold higher than non-Kuwaitis. However, due to the pandemic, despite them being younger, non-Kuwaitis experienced a much higher percentage of excess deaths. This emphasizes that migrant workers are particularly vulnerable to this infection and its social, economic and environmental consequences.

Yet, it is a similar story. Migrant workers in Kuwait had more than two-fold increase in the odds of requiring intensive care or dying from COVID-19 compared to Kuwaitis after adjustment for baseline characteristics including age and co-morbidities(15,20). Significant spreading and clustering outbreaks of COVID-19 in Kuwait were shown to be in areas populated by migrant workers due to large population density and cramped housing(16). A cumulative risk assessment of the effects of COVID-19 on migrant workers in Kuwait showed that stressors arising from domains other than the individual-level are inseparable from risks of adverse health(14). Migrant workers were subject to layoffs and furloughs, facing job uncertainty, defaulting on their rent and sometimes unable to meet daily needs. Additionally, migrant workers in Kuwait were also shown to be especially vulnerable to death from environmental exposures such as extreme heat, dust storms and air pollution(21,22). Disparities in the pandemic excess deaths now is just another confirmatory piece of evidence of structural health inequities that are based on nationality and migration status.

There are many reasons that may explain the global excess deaths phenomenon and the disparities associated with it during a public health crisis. First, the social and environmental determinants of health have been inversely associated with noncommunicable diseases (NCDs); those most deprived are more likely to suffer from NCDs than other more privileged individuals and communities(23,24). Thus, among other things, it is possible that ethnic and racial disparities in COVID-19 mortality and excess deaths are attributable to a higher prevalence of NCDs in marginalized communities. Second, delayed presentation –either due to personal fear of contracting the virus or difficult access to healthcare, is associated with more extensive disease and therefore requiring more aggressive intervention. For instance, a rise in surgical emergencies such as obstructed hernia with bowel ischemia, diabetic foot ulcers requiring amputation and appendicular abscess was noted during this pandemic(25). Another example, patients with oncological conditions presented with complications and that can be attributed to the temporary stop of elective surgeries(25). The same can be said regarding medical emergencies, it was observed that delayed presentation of myocardial infarction is associated with higher mortality and more complication such as left ventricular free wall rupture (which tends to be fatal) and necrosis(26,27). Furthermore, the worldwide postponement of medical care most likely resulted in a backlog of appointments, procedures and surgeries due to increase demand and limited healthcare facilities. This further delay is likely to have an adverse impact on population health(28).

We compared Kuwait’s percentage excess deaths of 56.0% (simple averaging) to a number of other countries from ***Our World In Data*** tracker of excess deaths(17). The method used to predict expected deaths for other countries in the repository included summing deaths over weeks or months. We were not able to do that for Kuwait given our restricted access to only annual deaths. Yet, at face value, excess deaths in Kuwait were still considerably greater than levels observed in hardly hit countries like the U.S. and Europe (**Supplemental Figure A**). For example, the 2020 annual percentage of excess deaths in the U.S., Spain and Italy are estimated to be 19, 18 and 15%, respectively. Compared to other Gulf states, excess deaths in Kuwait exceeded those from Oman (24%) and Qatar (14%). Comparable numbers from other Gulf countries like Saudi Arabia or the United Arab Emirates were not available. Kuwait’s numbers are however comparable to South American countries such as Ecuador (63%), Bolivia (52%) and Mexico (47%). Given that the pandemic may lead to fewer deaths from accidental causes, the excess numbers of deaths observed in Kuwait in 2020 are concerningly sizeable. It is acknowledged that COVID-19 mortality figures are underestimates of the actual death toll (29,30). By December 31 ^st^, 2020, the cumulative COVID-19 official reported mortality in Kuwait was 934 deaths (31). According to our estimates this leaves an additional 2400 overall unexplained deaths. COVID-19 deaths alone need to be underestimated by a factor of nearly 3.5 in order to explain the additional mortality. However, we argue that indirect fatalities from delayed and disrupted care due to cancer, circulatory and other causes may have played a significant role.

This study has a number of limitations. At the time of writing, data on gender, age or cause of death stratification were not available. Although we present two methods in estimating 2020 expected deaths, there are more advanced forecasting techniques in predicting expected mortality such as seasonal autoregressive integrated moving average (ARIMA) and machine learning. However, we were limited in our temporal resolution of annual deaths. Daily, weekly or monthly observed deaths were not publicly available. Because of that, the readers must be cautioned that our overall estimation of excess deaths percentage may be exaggerated. Finally, the disparity in migrant workers excess deaths is restricted to Kuwait and cannot be generalized to other contexts. Yet, the picture is unlikely to be completely different in other Gulf countries that are hosts to millions of migrant workers.

## Conclusion

Based on historical annual deaths in Kuwait, the mortality burden of COVID-19 pandemic is substantially higher than what the official tally might suggest. Systematically disadvantaged marginalized subpopulations like migrant workers are coming off worse and bearing a larger weight from the pandemic. Population health was severely and differentially impacted by COVID-19 warranting public health interventions that are sensitive to underlying structural and societal determinants that generate stark inequities and disparities.

## Data Availability

Data is made available in the supplemental material

https://www.paci.gov.kw/stat/TimeSeries.aspx

## Notes

### Competing Interest Statement

The authors have declared no competing interest.

### Funding Statement

No funding was obtained for this study

### Author Declarations

Analysis of aggregated publicly available data in which no IRB is required.

