## Supplemental for "Disparities in Excess Deaths from the COVID-19 Pandemic Among Migrant Workers in Kuwait"

*Alahmad et al.*

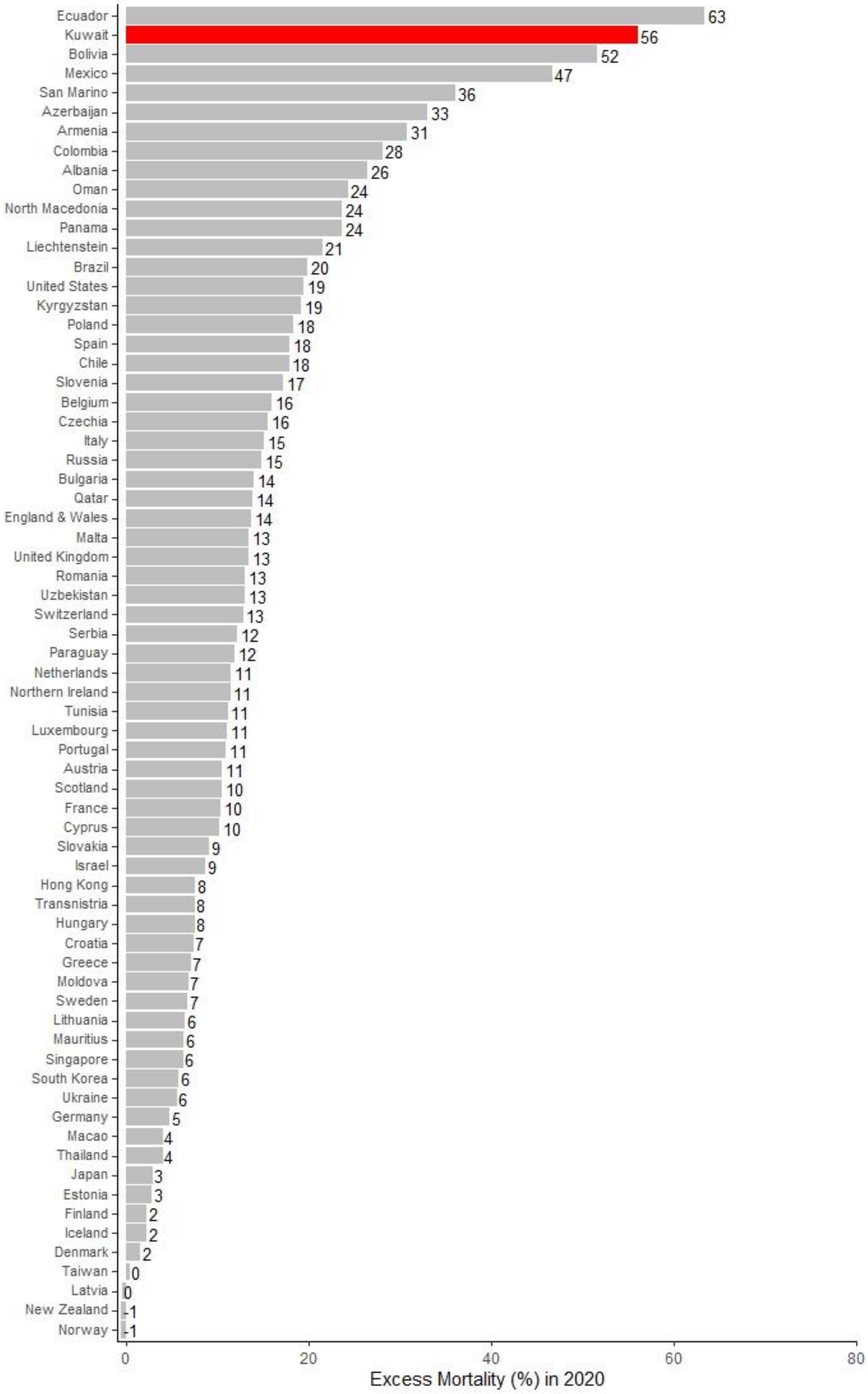
Ecuador 63
Kuwait 56
Bolivia 52
Mexico 47
San Marino 36
Azerbaijan 33
Armenia 31
Colombia 28
Albania 26
Oman 24
North Macedonia 24
Panama 24
Liechtenstein 21
Brazil 20
United States 19
Kyrgyzstan 19
Poland 18
Spain 18
Chile 18
Slovenia 17
Belgium 16
Czechia 16
Italy 15
Russia 15
Bulgaria 14
Qatar 14
England & Wales 14
Malta 13
United Kingdom 13
Romania 13
Uzbekistan 13
Switzerland 13
Serbia 12
Paraguay 12
Netherlands 11
Northern Ireland 11
Tunisia 11
Luxembourg 11
Portugal 11
Austria 11
Scotland 10
France 10
Cyprus 10
Slovakia 9
Israel 9
Hong Kong 8
Transnistria 8
Hungary 8
Croatia 7
Greece 7
Moldova 7
Sweden 7
Lithuania 6
Mauritius 6
Singapore 6
South Korea 6
Ukraine 6
Germany 5
Macao 4
Thailand 4
Japan 3
Estonia 3
Finland 2
Iceland 2
Denmark 2
Taiwan 0
Latvia 0
New Zealand -1
Norway -1
0
20
40
60
80
Excess Mortality (%) in 2020

**Supplemental Figure A:** Percentage of excess mortality in 2020 across different countries around the world. Only countries with full weekly (52 weeks) or monthly (12 months) data were selected from *Our World In Data* excess deaths tracker project. Expected deaths for each country was calculated using a 5-year average of deaths from 2015 to 2019. Observed and expected deaths were summed over weeks or months for each country, then excess deaths were calculated as (observed – expected)/expected*100 for the year 2020. The method for predicting Kuwait's expected deaths is explained in the main manuscript.

**Supplemental Raw data:** Deaths, nationality and population data from the Public Authority for Civil Information (PACI), Reported December 31st, 2020.

| year | deaths_PACI | nationality | pop |
|---|---|---|---|
| **2005** | 2348 | K | 992217 |
| **2005** | 2029 | NK | 1998972 |
| **2005** | 4377 | T | 2991189 |
| **2006** | 2608 | K | 1023316 |
| **2006** | 2237 | NK | 2159644 |
| **2006** | 4845 | T | 3182960 |
| **2007** | 2747 | K | 1054598 |
| **2007** | 2253 | NK | 2345039 |
| **2007** | 5000 | T | 3399637 |
| **2008** | 2741 | K | 1087552 |
| **2008** | 2253 | NK | 2354261 |
| **2008** | 4994 | T | 3441813 |
| **2009** | 2775 | K | 1118911 |
| **2009** | 2430 | NK | 2365970 |
| **2009** | 5205 | T | 3484881 |
| **2010** | 2798 | K | 1148363 |
| **2010** | 2358 | NK | 2433691 |
| **2010** | 5156 | T | 3582054 |
| **2011** | 2877 | K | 1183185 |
| **2011** | 2444 | NK | 2514107 |
| **2011** | 5321 | T | 3697292 |
| **2012** | 3070 | K | 1212436 |
| **2012** | 2504 | NK | 2611292 |
| **2012** | 5574 | T | 3823728 |
| **2013** | 3084 | K | 1242499 |
| **2013** | 2593 | NK | 2722645 |
| **2013** | 5677 | T | 3965144 |
| **2014** | 3186 | K | 1275857 |
| **2014** | 2633 | NK | 2816136 |
| **2014** | 5819 | T | 4091993 |
| **2015** | 3261 | K | 1307605 |
| **2015** | 2838 | NK | 2931401 |
| **2015** | 6099 | T | 4239006 |
| **2016** | 3439 | K | 1337693 |
| **2016** | 2809 | NK | 3073431 |
| **2016** | 6248 | T | 4411124 |
| **2017** | 3432 | K | 1370013 |
| **2017** | 2924 | NK | 3130463 |
| **2017** | 6356 | T | 4500476 |
| **2018** | 3433 | K | 1403113 |
| **2018** | 2941 | NK | 3218525 |
| **2018** | 6374 | T | 4621638 |
| **2019** | 3687 | K | 1432045 |
| **2019** | 3209 | NK | 3344362 |
| **2019** | 6896 | T | 4776407 |
| **2020** | 4756 | K | 1459970 |
| **2020** | 5219 | NK | 3210743 |
| **2020** | 9975 | T | 4670713 |

**Supplemental Table A:** Estimates of expected and excess deaths in 2020 stratified by nationality using historical mortality data (2001 to 2018) from Kuwait's Central Statistical Bureau, and Kuwait's Public Authority for Civil Information deaths for the 2019 to 2020 period.

| | Total | Kuwaitis | Non-Kuwaitis |
|---|---|---|---|
| **Observed Deaths in 2020** | 9975 | 4756 | 5219 |
| **Regression Estimate:** | | | |
| *Expected Deaths (95% CI)* | 6680 (6422 to 6948) | 3464 (3330 to 3603) | 3216 (3092 to 3345) |
| *Excess Deaths (95% CI)* | 3295 (3027 to 3553) | 1292 (1153 to 1426) | 2003 (1874 to 2127) |
| *Excess Deaths % (95% CI)* | 49.3 (43.6 to 55.3) | 37.3 (32.0 to 42.8) | 62.3 (56.0 to 68.8) |
| **Simple Averaging Estimate:** | | | |
| *Expected Deaths (95% CI)* | 6640 (6480 to 6800) | 3471 (3355 to 3586) | 3169 (3059 to 3280) |
| *Excess Deaths (95% CI)* | 3335 (3175 to 3495) | 1285 (1170 to 1401) | 2050 (1939 to 2160) |
| *Excess Deaths % (95% CI)* | 50.2 (46.7 to 53.9) | 37.0 (32.6 to 41.7) | 64.7 (59.1 to 70.6) |
